# Lifetime risk of incident dementia and incident mild cognitive impairment in older adults

**DOI:** 10.1101/2025.08.29.25334749

**Authors:** Lianlian Du, Lei Yu, Tianhao Wang, Patricia A. Boyle, Lisa L. Barnes, David X. Marquez, David A. Bennett

## Abstract

**Background and Objectives:** To estimate the lifetime risk of dementia and mild cognitive impairment (MCI) from age 55 to 105, accounting for competing risk of death, and to examine differences by sex and race.

**Methods:** We analyzed data from five harmonized longitudinal cohort studies at the Rush Alzheimer’s Disease Center, including 4611 community dwelling older adults for lifetime dementia risk estimation and 3915 for lifetime MCI risk estimation. Incident dementia and MCI were identified through annual clinical evaluations.

Nonparametric cumulative incidence function curves estimated lifetime risk, adjusting for competing risk of death and left truncation. Additional analyses assessed lifetime risk from index ages 55, 65, 75, and 85 and examined differences by sex and race.

**Results:** The lifetime risk of incident dementia after age 55 was 43% (95% CI: 38–47), with a median age at diagnosis of 88 years(IQR: 83–92). For MCI, the lifetime risk was 62% (95% CI: 57–67), with a median age at diagnosis of 86 years(IQR: 80–90). Females had higher lifetime risks than males for both dementia (45% vs. 39%) and MCI (63% vs. 60%). Racial differences were smaller for dementia (45% in Black vs. 44% in White participants). For MCI, Black adults had higher lifetime risk before age 90.

**Discussion:** These findings extend dementia lifetime risk estimation beyond age 90 among diverse older adults to provide lifetime risk estimates for MCI while accounting for the competing risk of death, highlighting the importance of prevention, and equitable public health strategies to reduce the burden of cognitive impairment.

## INTRODUCTION

Dementia is a growing public health crisis, affecting over 7 million individuals in the United States, and this number is projected to reach about 14 million by 2060.^1,2^ Among individuals aged 90–95, dementia reportedly contributes to more than 40% of deaths in females and 30% in males.^3^ The financial impact is severe, with healthcare and long-term care costs estimated to reach $360 billion in 2024, in addition to about $350 billion in unpaid caregiving costs.^4^ This socioeconomic burden is expected to grow steeply with ageing populations and continuing increases in life-expectancy worldwide. Mild cognitive impairment (MCI) is also a serious concern, as it links to an increased risk of mortality and dementia.^4^ In the U.S., individuals with MCI are projected to grow from more than 12 million in 2020 to about 22 million in 2060.^1^ Prevalence of MCI increases with age, affecting 7% of those aged 60–64 and 25% of those aged 80–84.^5^ Since no cure exists for dementia, prevention is critical.^6^

The lifetime risk of dementia is a critical public health measure that informs public policy, enhances engagement by other stakeholders (e.g., American Heart Association and the Alzheimer’s Association), and helps estimate the potential benefits of early interventions.^4,7,8^ Estimating lifetime risk is challenging due to competing mortality risk and varying ages at study entry. We previously reported that death following incident dementia occurred on average less than 3 years among those over age 85.^9^ Thus, capturing dementia incidence in the oldest old requires assessments at relatively short intervals to identify cases that emerge and progress between visits. Further, standard methods like the Kaplan-Meier estimator often overestimate risk by failing to account for competing events; this is especially problematic for late-life diseases like dementia for which competing mortality is high.^10^ To avoid this bias, lifetime estimates should incorporate both competing mortality and adjustment for left truncation, i.e., age at study entry.^11^ Given the known sex, racial, and ethnic differences in dementia risk, estimating lifetime risk across subgroups is essential.^4,7,12^ Prior research has made significant advances in documenting the risk and burden of cognitive impairment.^6,12–15^ A recent study using a subset of these data predicted dynamic lifetime risk of Alzheimer’s disease using longitudinal cognitive assessment data.^16^ Further, racial disparities in lifetime risk remain poorly characterized, as most studies focus on non-Latino White populations.^12,18^ In addition, few studies include large numbers of persons age 90+. Finally, to our knowledge, only one study estimated MCI lifetime risk.^13^ Given the growing societal burden of cognitive impairment^2^ and the increasing complexity of daily life in a technology-driven world,^19^ accurately estimating its lifetime risk is crucial.

Here, we leveraged longitudinal data from five harmonized cohort studies of aging and dementia from the Rush Alzheimer’s Disease Center (RADC) to estimate the lifetime risks of dementia and MCI. Further, we investigated potential differences in the lifetime risks by sex and race. These analyses highlight overall risks and disparities in cognitive impairment and dementia incidence across aging populations.

## METHODS

### Participants

The study participants came from one of five ongoing cohort studies of aging and dementia at the RADC: the Religious Orders Study (ROS)^20^, the Rush Memory and Aging Project (MAP)^20^, the Minority Aging Research Study (MARS)^21^ the African American Clinical Core (AACORE)^22^ and the Latino CORE (LATC).^23^ ROS began in 1994 and enrolls older Catholic priests, nuns, and brothers throughout the United States. MAP started in 1997 and enrolls older adults from retirement communities and subsidized senior housing facilities and individual homes throughout northeastern Illinois. MARS and AACORE were initiated in 2004 and 2008, respectively, and enroll older African Americans. LATC, established in 2015, enrolls older Latino Americans. Participants of MAP, MARS, AACORE and LATC all come from the same geographical area in the midwest. To facilitate combined analyses, the five studies share similar protocols for study design and data collection, as outlined previously.^23^ Participants enroll without known dementia. All participants are followed annually until death. Individual studies were approved by an institutional review board at Rush University Medical Center, and all participants provided written informed consent. For lifetime dementia risk estimation, we excluded participants with a diagnosis of dementia at baseline (n = 281), those without follow-up data (n = 248), including individuals who were not yet eligible for their annual evaluation at the time of these analyses (2025 January), died, or declined further clinical participation. We also excluded participants younger than 55 years at baseline (n = 8) to allow for proper left truncation modeling. To ensure diagnostic consistency, we further excluded deceased participants if the interval between their last visit and death exceeded three years (n = 194; see *Dementia and MCI Ascertainment*), resulting in a final analytic sample of 4611 participants. For lifetime MCI risk estimation, we applied the same exclusions as above and further removed participants who had MCI at baseline (n=696), yielding a final analytic sample of 3915 participants. See Supplementary Figure 1 (STROBE flow diagram) for details.

### Dementia and MCI ascertainment

The five RADC cohort studies share a uniform structured clinical evaluation, as described elsewhere.^23–25^ Briefly, at baseline and annual follow-ups, each evaluation includes a medical history, neurological examination, and neuropsychological assessment. Diagnoses follow a three-stage process, as previously reported.^24–26^ Dementias uses the criteria established by the joint working group of the National Institute of Neurological and Communicative Disorders and Stroke and the Alzheimer’s Disease and Related Disorders Association (NINCDS/ADRDA) for the clinical diagnosis of dementia and clinical Alzheimer’s dementia.^27^ Individuals are classified as MCI if they exhibit cognitive impairment based on the neuropsychologist’s assessment but did not meet dementia criteria. Importantly, each assessment each year was made without access to prior diagnoses or other data from prior years.

Dementia was treated as an all-absorbing state with onset defined as the first recorded diagnosis. For MCI, we applied a conservative definition requiring at least two consecutive data points, with onset defined as the first MCI diagnosis followed by either a subsequent MCI diagnosis, dementia diagnosis, or death within three years. Importantly, cognitive scores in the MCI–Death group were not significantly different from the MCI– MCI group, after adjusting for age, sex, race & ethnicity and education, suggesting that inclusion of MCI-Death into the MCI category is reasonable. More information on group cognitive scores is presented in Supplementary materials(Supplementary Figure 2).

### Demographic characteristics

All participants report their sex (i.e., male or female), date of birth, and years of education. Age is calculated from birth date to date of the clinical examination. Participants also report their race (e.g. African American/Black) and ethnicity (e.g. Latino/Hispanic: yes or no) according to the 1990 US Census questions.

### Statistical Analysis

We examined baseline characteristics and the proportions of dementia and MCI cases. Differences in demographics and incidence rates between subgroups by sex and race were assessed using chi-squared tests, Fisher’s exact tests, analysis of variance (ANOVA), or Kruskal-Wallis tests, as appropriate.

We estimated the lifetime risk of dementia and MCI using non-parametric cumulative incidence function curves from ages 55 to 105, while accounting for competing risks to prevent overestimation of lifetime risks.

A widely used approach for competing risks analysis is the proportional subdistribution hazards model by Fine and Gray.^28^ Given our older baseline age, we used age as the timescale and accounted for left truncation. However, because implementing the Fine-Gray method with left truncation can be challenging, we applied a generalized linear model with pseudo-observations of the Aalen-Johansen estimator, ^29–31^ which accounts for left truncation and has been shown to produce minimal bias.^11^

We defined two competing events ɛ_l_: where ɛ=1is the diagnosis of interest (dementia or MCI) and ɛ=2 is the competing event (death without diagnosis). Using age as the time scale, we defined *L* as the entry age, *T* as the age at failure, *C* as the age at censoring, and an indicator *δ*= 1(r< *C*). We considered an index age τ_0_ such that *L*≥τ_0_. The observed data for each individual *i*(*i*=1,…, *n*) consisted of (*L*_l_, *X*_l_,δ_l_ ·ɛ_l_,*z*_l_), where we observe *X*=*T^C* represents the age at diagnosis, death without diagnosis, or censoring, and *z* denotes a matrix of covariates (e.g., sex). Lifetime risk was estimated as the cumulative incidence from index age τ_0_ (e.g., 55 years) to an advanced age τ (e.g., 105 years):

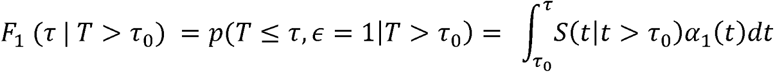

where α_1_ (t) is the cause-specific hazard function for event 1, and 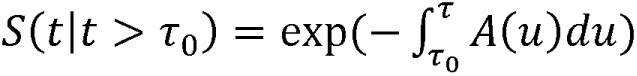 is the overall survival function where *A*(*t*) is the cumulative all-cause hazard function. Lifetime risk was estimated non-parametrically using the Aalen-Johansen estimator, accounting for left truncation. Details on the pseudo-observation model have been previously described.^11^ For this study, age 55 was used as the index age, with entry age defined as the age at baseline. Person-time was calculated from age 55 until dementia diagnosis, death free of dementia, administrative censoring (January 2025) or age 105, whichever occurred first.

In secondary analyses, we estimated lifetime risk using cumulative incidence function curves with index ages at 65, 75, and separately 85 years. Among individuals with incident dementia, we calculated the median age at diagnosis and the proportion diagnosed between ages 55–74, 75–84, 85–95, and 95–105. We then repeated the analysis for MCI, and then stratified by sex and race.

All analyses were conducted using R (version 4.4.1), with statistical significance determined at α=0.05.

## RESULTS

### Baseline Characteristics and Incidence of Dementia and MCI

Baseline characteristics of participants in the dementia (n = 4611; 74.4% female, 27.5% Black; 6.4% Latino among White, 2.0% Latino among Black) and MCI (n = 3915; 75.8% female, 27.6% Black; 6.7% Latino among White, 1.8% Latino among Black) lifetime risk sets are summarized in Table 1. Baseline age distributions (55–64, 65–74, 75–84, 85+) were 206, 1799, 1904, and 702 for the dementia set, and 190, 1649, 1573, and 503 for the MCI set. Differences in demographic characteristics between the two samples are shown in Table 1, with further subgroup comparisons by sex and race in Supplementary Tables 1 and 2.

**Table 1.** Characteristics of Study Participants in Estimating Lifetime Risk of Dementia and MCI.

| Characteristic | N = 4611 | N = 3915 |
| --- | --- | --- |
| Age at baseline, years | 76.59 (7.72) | 75.87 (7.53) |
| Age at last visit, years | 85.36 (7.87) | 85.08 (7.92) |
| Follow up years | 8.50 (5.76) | 8.92 (5.83) |
| Female, n (%) | 3431 (74.4) | 2967 (75.8) |
| White, n (%) | 3151 (68.4) | 2661 (68.0) |
| Education, years | 15.77 (3.95) | 15.83 (3.98) |
| *APOE* ε4 carriers, n (%) | 990 (26.7)<sup>a</sup> | 795 (25.4)<sup>b</sup> |
| Study, n (%) |  |  |
| ROS | 1323 (28.7) | 1144 (29.2) |
| MAP | 1962 (42.6) | 1600 (40.9) |
| MARS | 748 (16.2) | 649 (16.6) |
| AA | 341 ( 7.4) | 309 ( 7.9) |
| LATC | 237 ( 5.1) | 213 ( 5.4) |
| Alive free of dementia/MCI, n (%) | 1872 (40.6) | 1575 (40.2) |
| Incident cases of dementia/MCI, n (%) | 1278 (27.7) | 1520 (38.8) |
| Deaths without dementia/MCI, n (%) | 1461 (31.7) | 820 (20.9) |
| Age at death | 86.82 (7.34) | 85.77 (7.60) |
| Interval between age at last and at death<br>(median [IQR]) | 0.69 [0.39, 1.01] | 0.69 [0.39, 1.04] |

Column 2 is the dataset in estimating the lifetime risk of Dementia, and column 3 is the dataset in estimating the lifetime risk of MCI. Values are mean (SD) and for the entire analytic sample unless otherwise stated.
Abbreviations: MCI, Mild Cognitive Impairment; ROS, Religious Orders Study; MAP, Memory and Aging Project; MARS, Minority Aging Research Study; AACORE, Clinical Core; LATC, Latino Core; *APOE*, Apolipoprotein E; IQR: Interquartile range
<sup>a</sup> Data missing for 901 participants; <sup>b</sup> Data missing for 790 participants.

**Table 2.** Median age at dementia and MCI diagnosis and distribution of diagnosis age, overall and by sex and race.

|  | Median age (IQR) at diagnosis, years | Percentage diagnosed between ages 55 and 74 | Percentage diagnosed between ages 75 and 84 | Percentage diagnosed between ages 85 and 95 | Percentage diagnosed between ages 95 and 105 |
| --- | --- | --- | --- | --- | --- |
| **Dementia** |  |  |  |  |  |
| Overall | 87.59 [82.65, 91.78] | 5.3% | 30.0% | 53.2% | 11.4% |
| Sex |  |  |  |  |  |
| Female | 87.94 [82.78, 92.37] | 5.7% | 27.6% | 54.1% | 12.6% |
| Male | 86.45 [82.28, 90.57] | 4.3% | 37.1% | 50.6% | 8.0% |
| Race |  |  |  |  |  |
| White | 88.53 [84.04, 92.68] | 3.0% | 26.2% | 57.1% | 13.6% |
| Black | 84.35 [80.01, 88.62] | 10.8% | 43.7% | 41.4% | 4.1% |
| **MCI** |  |  |  |  |  |
| Overall | 85.61 [80.12, 90.13] | 9.7% | 37.6% | 46.1% | 6.6% |
| Sex |  |  |  |  |  |
| Female | 85.90 [80.26, 90.45] | 9.8 % | 35.4% | 47.4% | 7.3% |
| Male | 84.47 [79.84, 89.52] | 9.3% | 44.1% | 42.0% | 4.6% |
| Race |  |  |  |  |  |
| White | 86.89 [82.22, 90.90] | 5.3% | 34.3% | 52.0% | 8.4% |
| Black | 80.55 [75.33, 85.67] | 22.1% | 48.8% | 27.9% | 1.2% |

Percentages may not add up to 100% due to rounding. IQR, interquartile range.

Importantly, age at baseline was similar for males and females, but Black adults were about five years younger among those without dementia as well as without cognitive impairment.

Among the dementia lifetime risk set, over a mean follow-up of 9 years (range: 1–29 years), there were 1278 incident dementia cases, 1461 deaths without dementia, and 1872 individuals alive and dementia-free at last visit. The total numbers of participants aged 55–74, 75–84, 85–94, and 95–105 years were 544, 1692, 1986, and 389. The distributions of the three outcomes: censor (alive and dementia-free), event (incident dementia), and competing risk (death without dementia) across different age groups are presented in Supplementary Figure 3A, and stratified by sex and race in Supplementary Figures 3B and 3C. Dementia incidence was higher in White participants compared to Black participants (31.3% vs. 21.2%; p < 0.001), but similar across sex (27.7% vs. 27.6%). Conversely, death without dementia was more common among White individuals than Black individuals (36.2% vs. 23.7%; p < 0.001) and more frequent among males than females (39.9% vs. 28.9%; p < 0.001). On average, Black participants were about six years younger at death (82 vs. 88).

Among participants in the MCI lifetime risk set, there were 1520 incident MCI cases, 820 deaths without MCI, and 1575 individuals alive and MCI-free at last visit. The total numbers of participants aged 55–74, 75– 84, 85–94, and 95–105 years were 567, 1584, 1541, and 223, respectively. The distributions of the three outcomes (alive and MCI-free, incident MCI, and death without MCI) across these age groups are presented in Supplementary Figure 4A. Stratified distributions by sex and race are shown in Supplementary Figures 4B and 4C, respectively. MCI incidence was higher among White participants (43.5% vs. 30.2%; p < 0.001), but similar by sex (40.9% in males vs. 38.2% in females). Death without MCI was more common in White participants (23.7% vs. 16.7%; p < 0.001) and male participants (26.3% vs. 19.2%; p < 0.001). On average, Black participants died six years earlier than White participants (81 vs. 87).

## Overall Results

### Lifetime Risk of Dementia

Starting at age 55, the lifetime risk of incident dementia up to age 105 was 43% (95% CI: 38–47) (Figure 1A). The cumulative incidence remained low, 6% by age 75 and increased sharply afterward. Interestingly, the increase slows and plateaus at 39% after age 95 and only increases to 43% over the next decade. The cumulative incidence of dementia at each year of age is shown in Supplementary Table 3. Among the 1278 incident dementia cases, the median age at diagnosis was 88 years (IQR: 83–92) (Table 2). Notably, 83% of cases were diagnosed between ages 75 and 95, with 53% occurring between ages 85 and 95.

**Figure 1.**
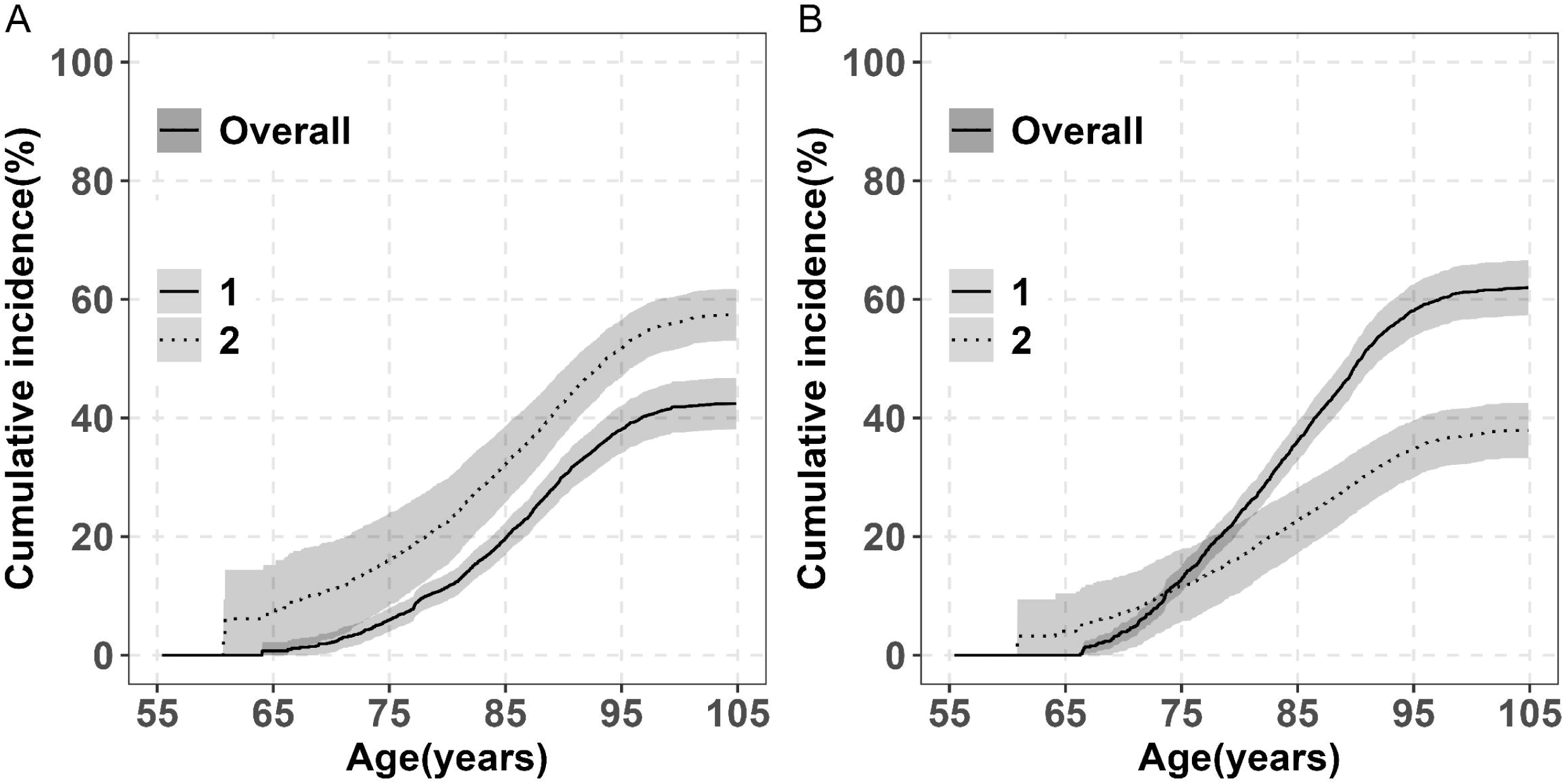
Overall Lifetime Risk of Dementia, MCI, and Death Without Diagnosis (Ages 55–105) in the Five RUSH Cohorts. Panel A: cumulative incidence of dementia (solid line) and death without dementia (dashed line) (n = 4,611). Panel B: cumulative incidence of MCI (solid line) and death without MCI (dashed line) (n = 3,915). Estimates are percentages accounting for competing risk of death. Shaded areas show 95% confidence intervals (CIs). Abbreviation: MCI, Mild Cognitive Impairment.

Lifetime risk was related to index ages (Table 3). Among participants who are alive and dementia-free, it rose from 43% among persons starting at age 55 to 48% among those starting at age 75. However, it was 45% among those starting at 85, likely due to a smaller sample size and shorter life expectancy, as suggested by the wider confidence interval.

**Table 3.** Lifetime risk of dementia and MCI after select index ages (ages 55, 65, 75, and 85) to 105Dyears, overall and by sex and race using primary analysis.

|  | Index age |  |  |  |
| --- | --- | --- | --- | --- |
|  | 55 years | 65 years | 75 years | 85 years |
| **Lifetime risk of dementia** | (n=4611) | (n=4405) | (n=2606) | (n=702) |
| Overall | 43 (38,47) | 45 (43,47) | 48 (46,51) | 45 (41,50) |
| Sex |  |  |  |  |
| Female | 45 (40,50) | 49 (46,51) | 49 (46,52) | 46 (40,51) |
| Male | 39 (35,43) | 38 (33,42) | 48 (42,54) | 44 (35,53) |
| Race |  |  |  |  |
| White | 44 (37,50) | 45 (42,48) | 48 (45,51) | 46 (42,51) |
| Black | 45 (40,50) | 47 (42,52) | 50 (43,56) | 35 (21,49) |
| **Lifetime risk of MCI** | (n=3915) | (n=3725) | (n=2076) | (n=503) |
| Overall | 62 (57,67) | 64 (62,67) | 69 (66,72) | 64 (59,69) |
| Sex |  |  |  |  |
| Female | 63 (58,69) | 67 (65,70) | 70 (67,73) | 64 (58,70) |
| Male | 60 (55,64) | 58 (52,63) | 67 (62,72) | 62 (51,73) |
| Race |  |  |  |  |
| White | 65 (62,67) | 64 (62,67) | 68 (65,71) | 65 (60,70) |
| Black | 60 (54,65) | 62 (57,67) | 69 (63,75) | 52 (34,70) |

Estimates are reported as percentages and indicate the cumulative incidence at the age of last observation (up to age 105 □ years) after accounting for the competing risk of death. The 95% CIs are reported in parentheses.

To account for competing risks, we estimated the cumulative incidence of death without dementia. Between ages 55 and 105, the overall cumulative incidence was 57% (95% CI: 53–62), as shown in Figure 1A. In parallel with the incidence of dementia, risk of death also plateaued at 51% at age 95 increasing to only 57% over the next decade. The cumulative risks of death at each year of age are provided in Supplementary Table 4.

### Lifetime Risk of MCI

Starting at age 55, the estimated lifetime risk of incident MCI up to age 105 was 62% (95% CI: 57–67), approximately 20% higher than the lifetime risk of incident dementia (Figure 1B). The cumulative incidence of MCI increased rapidly after age 65, roughly 10 years earlier than for dementia. In parallel with dementia, the increase slows and plateaus at 58% after age 95 and only increases to 62% over the next decade.

Corresponding cumulative incidence estimates across ages are provided in Supplementary Table 3. Among 1520 incident MCI individuals, the median age at diagnosis was 86 years (IQR: 80–90 years), about two years younger than the median age at dementia diagnosis (Table 2). 84% of MCI cases were diagnosed between ages 75 and 95, with 46% occurring between ages 85 and 95.

Lifetime risk of MCI was associated with index age (Table 3). Among participants who are alive and dementia-free, it rose from 62% among persons age 55 to 69% among those age 75. Among those age 85, it declined slightly to 64%, likely reflecting smaller sample sizes and shorter life expectancy, as indicated by wider confidence intervals.

The overall cumulative incidence of death without MCI between ages 55 and 95 was 35% increasing to only 38% (95% CI: 33–43) at age 105 (Figure 1B). Age-specific cumulative incidence estimates corresponding to this figure are detailed in Supplementary Table 4.

### Sex Differences in Lifetime Risk Lifetime Risk of Dementia by Sex

Females had a higher lifetime risk of dementia than males (45% vs. 39%) (Figure 2A), with detailed estimates provided in Supplementary Table 3. Most of the differences were seen after age 90. The increasing risk of dementia attenuated after age 95 for both males and females. The median age at diagnosis was approximately two years later in females compared to males (88 vs. 86, p < 0.01; Table 2). While most dementia diagnoses occurred after age 75 in both sexes, a greater proportion of females were diagnosed between ages 85 and 105 (67% vs. 59%), whereas males had a higher proportion of diagnoses between ages 75 and 84 (37% vs 28%).

**Figure 2.**
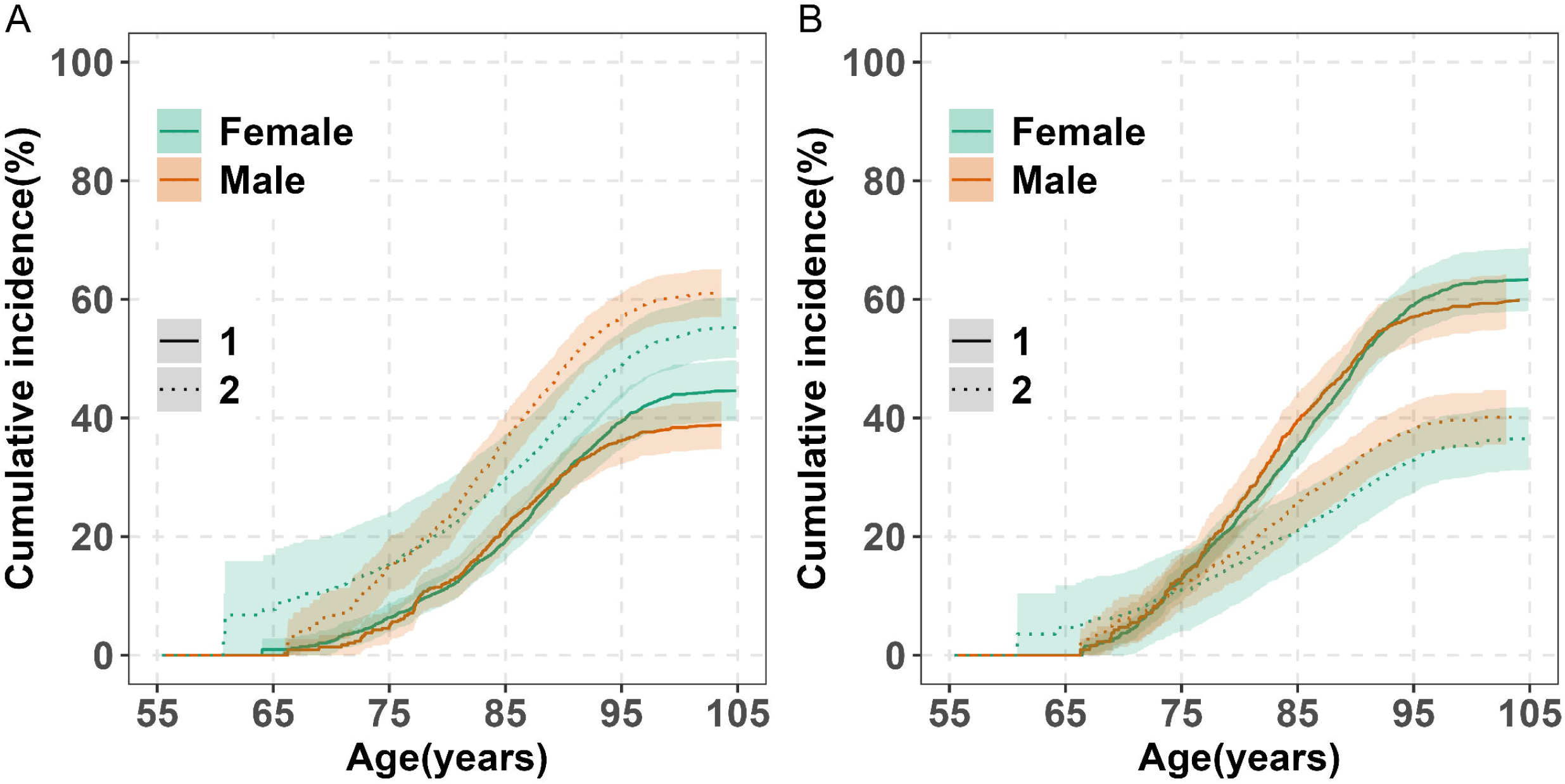
Lifetime Risk of Dementia, MCI, and Death Without Diagnosis by Sex (Ages 55–105) in the Five RUSH Cohorts. Panel A: The cumulative incidence of dementia (solid lines) and death without dementia (dashed lines) is shown for female and male (colored separately). Panel B: The cumulative incidence of MCI (solid lines) and death without MCI (dashed lines) is also shown for female and male. Estimates are percentages accounting for the competing risk of death. Shaded areas show 95% confidence intervals (CIs). Abbreviation: MCI, Mild Cognitive Impairment.

Lifetime risk was related to index ages for both sexes (Table 3). Among participants alive and dementia-free, it increased from 45% in females and 39% in males if estimates started at age 55 to 49% and 48%, respectively, if started at age 75, but it lowered slightly to 46% in females and 44% in males if started at age 85, likely due to smaller sample sizes and shorter life expectancy, as indicated by wider confidence intervals. A notable sex difference was observed at age 65, with lifetime risk at 49% in females and 38% in males.

The cumulative incidence of death without dementia from age 55 to 105 was higher in males than in females (61% vs. 55%) with the differences starting at about age 80 (Figure 2A; Supplementary Table 4).

### Lifetime Risk of MCI by Sex

The lifetime risk of MCI was higher in females than males through age 105 (63% vs. 60%) (Figure 2B; Supplementary Table 3), with most differences emerging after age 90. MCI risk plateaued after age 95 for both males and females. Like dementia, females were diagnosed with MCI approximately two years later than males (median age: 86 vs. 84 years, p < 0.05; Table 2). Most MCI cases occurred after age 75 in both sexes, with a higher proportion of diagnoses between ages 85 and 105 in females (54% vs. 47%), and between ages 75 and 84 in males (44% vs 35%).

Lifetime MCI risk was related to index ages for both sexes (Table 3). Among participants alive and MCI-free, it increased from 63% for females, 60% for males among persons starting at 55 to 70% for females, and 67% for males among those starting at age 75. However, it was 64% for females, 62% for males among those starting at 85, likely due to a smaller sample size and shorter life expectancy, as suggested by the wider confidence intervals.

The cumulative incidence of death without MCI from age 55 to 105 was higher in males than in females (40% vs. 37%) (Figure 2B and Supplementary Table 4).

### Racial Differences in Lifetime Risk Lifetime Risk of Dementia by Race

Starting at age 55, the lifetime risk of dementia through age 105 was slightly higher in Black than White participants (45% vs. 44%). It is important to note that projections for Black adults extended only to age 100 due to their younger baseline ages and ages at death (Figure 3A; Supplementary Table 3). Dementia was diagnosed approximately five years earlier in Black individuals compared to White individuals (median age: 84 vs. 89 years, p < 0.001; Table 2). A greater proportion of dementia cases in Black adults occurred before age 75 (11% vs. 3%) and between ages 75 and 84 (44% vs. 26%), compared to White participants.

**Figure 3.**
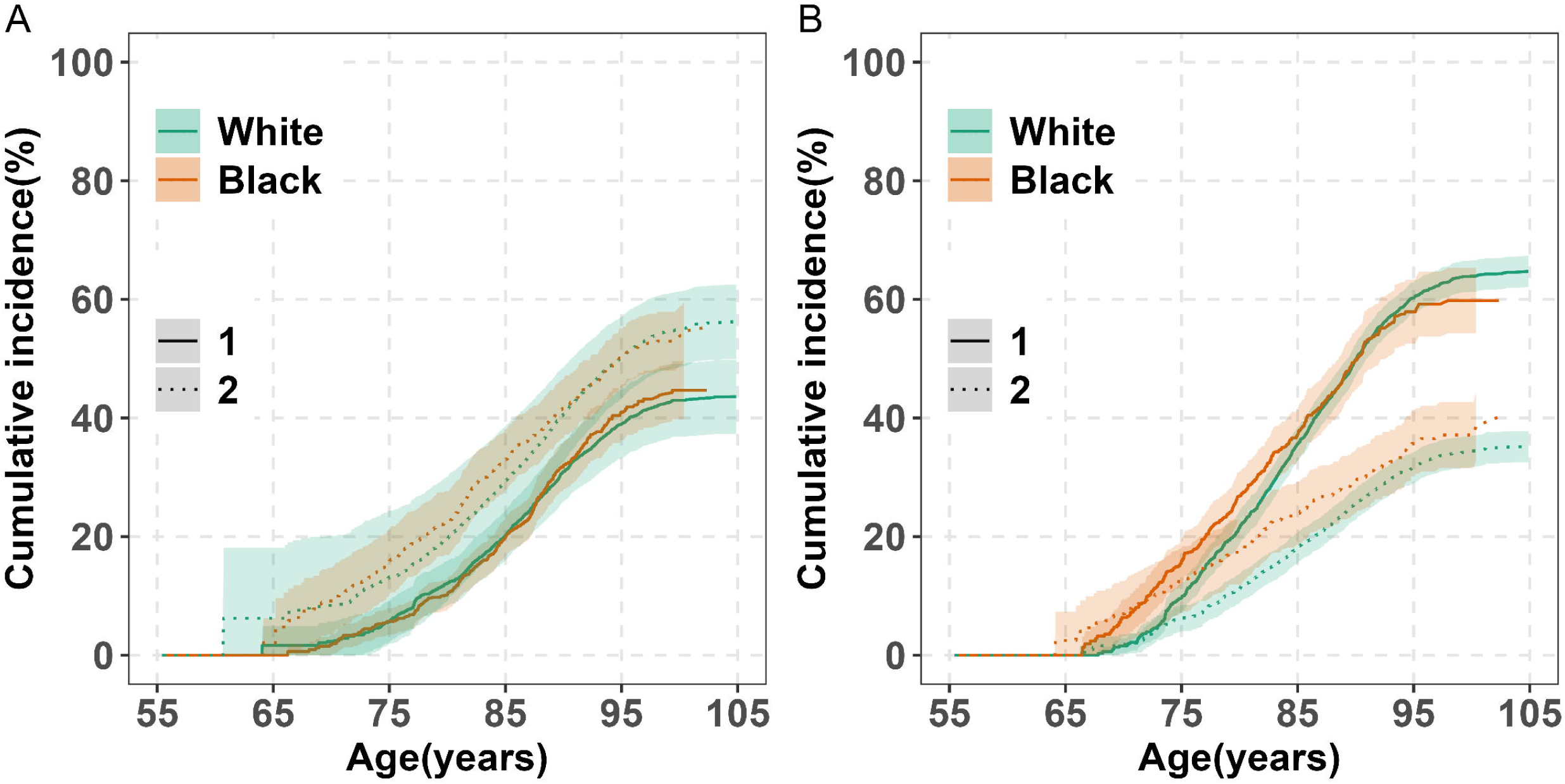
Lifetime Risk of Dementia and MCI and Death without Diagnosis by Race (Ages 55–105) in the Five RUSH Cohorts. Panel A: The cumulative incidence of dementia (solid lines) and death without dementia (dashed lines) is shown for white and black (colored separately). Panel B: The cumulative incidence of MCI (solid lines) and death without MCI (dashed lines) is also shown for white and black. Estimates are percentages accounting for the competing risk of death. Shaded areas show 95% confidence intervals (CIs). Abbreviation: MCI, Mild Cognitive Impairment.

Lifetime risk was associated with index age for both racial groups (Table 3). Among participants alive and dementia-free, it rose from 44% in White adults and 45% in Black adults among persons aged 55 to 48% and 50%, respectively, among those age 75. Among those age 85, it declined to 46% in White adults and 35% in Black adults, likely reflecting smaller sample sizes and shorter life expectancy, as suggested by wider confidence intervals.

Black individuals had a higher cumulative incidence of death without dementia before age 85 (from ages 55 to 85, 33% vs. 29%; Figure 3A; Supplementary Table 4) with the differences starting at about age 70.

However, at older ages, racial differences diminished with a modest cross-over at about age 95 driven by females, a phenomenon reported previously.^32^

### Lifetime Risk of MCI by Race

Starting at index age 55, Black adults had a higher lifetime risk of MCI beginning around age 65, but this risk was lower after age 95 compared to White adults. From ages 55 to 75, the estimated risk was 16% in Black participants versus 10% in White participants, and from ages 55 to 105, 60% versus 65% (Figure 3B; Supplementary Table 3). Projections for Black participants extended only to age 100 due to younger baseline ages and shorter life expectancy, with wider confidence intervals indicating greater uncertainty at older ages. MCI diagnosis occurred approximately six years earlier in Black than in White participants (median age: 81 vs. 87 years, p < 0.001; Table 2). A greater proportion of MCI diagnoses in Black participants occurred before age 75 (22% vs. 5%) and between ages 75 and 84 (49% vs. 34%).

Lifetime MCI risk was associated with index age in both racial groups (Table 3). Among participants alive and MCI-free, it rose from 65% in White adults and 60% in Black adults among persons starting at 55 to 68% and 69%, respectively, among those starting at 75. However, it was 65% for White adults, 52% for White adults among those starting at 85, likely due to a smaller sample size and shorter life expectancy, as indicated by the wider confidence interval.

Death without MCI was more frequent in Black adults, with a cumulative incidence through age 105 of 39% compared to 35% in White adults (Figure 3B and Supplementary Table 4); differences became apparent beginning around age 65.

## DISCUSSION

This study provides estimates of the lifetime risk of dementia and mild cognitive impairment (MCI) from age 55 to 105 using high-quality data from five harmonized longitudinal cohorts. By incorporating competing risk and left truncation methods, our findings build upon previous estimates. In the absence of effective public health strategies to reduce dementia incidence, our findings suggest that nearly half of older adults will develop dementia, while close to 2/3rds will develop MCI during their lifetime, with notable differences across sex and race. Actual numbers may be lower as some evidence suggests a slight decline in age-specific dementia prevalence over the past four decades,^33^ likely reflecting improvements in education, cardiovascular health, and lifestyle. However, rising life expectancy, and persistent racial differences in healthcare access may offset these gains.^34^ While the figures suggest that dementia incidence plateaus in the oldest old, the analytic approach precludes drawing this inference. Prior studies reported that dementia incidence increases with age but may peak and decline among the extremely old,^35^ or rise at a slower rate beyond age 85,^36^ and our previous work similarly demonstrated that pathologic AD peaked around age 95 and leveled off thereafter.^37^ However, not all findings are consistent and whether dementia incidence truly declines with advancing age remains uncertain.^38–40^

### Lifetime Risk Compared with Previous Studies

Our most striking finding is the high lifetime burden of MCI which begins to rise as early as age 65, nearly a decade before dementia onset. This underscores a large window for intervention and the urgent need to prioritize MCI as a target for early detection and prevention of dementia. This study is the first to our knowledge to estimate cumulative MCI risk from ages 55–105, providing novel insights into the progression and public health burden of MCI. A previous study using Health and Retirement Study (HRS) data reported higher MCI lifetime risk (71% for females and 61% for males),^13^ likely due to broader diagnostic criteria and younger participant age. By contrast, our use of a more conservative definition, requiring two consecutive assessments, reduced misclassification but resulted in later diagnosis ages and lower risk estimates.

We also observed higher lifetime risk of dementia, with most cases occurring between ages 75 and 95, and the risk increased progressively with older index ages 65 and 75, further supporting the importance of long-term cognitive surveillance in aging populations. Compared to previous studies, our dementia risk estimates are consistent or higher,^7,12,13,15,17,41^ which may reflect differences in population age distribution, diagnostic methods, follow-up frequency, or inclusion of left truncation. For example, our findings extend those of the ARIC study^12^ by estimating risk beyond age 95 and reveal slightly higher risk prior to age 84. Similarly, compared to the HRS,^13^ our dementia estimates were higher but with later onset ages. These differences likely are due to variations in study design, diagnostic criteria, and competing risk adjustments.

### Sex Differences in Lifetime Risk

Our findings did not show significant age-specific sex differences in the incidence of dementia or MCI, but females had a greater lifetime burden of both dementia and MCI compared to males, largely driven by higher mortality among males prior to dementia and MCI onset. This pattern aligns with multiple prior studies reporting consistently higher lifetime dementia risk in females, including ARIC (48% vs. 35%),^12^ two Rotterdam studies (31% vs. 19% and 33% vs. 16%),^8,42^ Framingham Heart Study^7,15^ (23% vs. 14%), HRS (37% vs. 24%),^13^ the Aging Demographics and Memory Study (ADAMS) (35% vs. 27%),^43^ and a biomarker-based study (27% vs. 21% at age 75).^16^ Our estimates were similar to ARIC and generally higher than others, which may reflect differences in population age structures, follow-up time, mortality rates, and diagnostic approaches. By using conservative clinical criteria and accounting for both competing risks and left truncation, our study provides more refined sex-specific lifetime risk estimates. The greater late-life dementia risk in females highlights the importance of sex-specific prevention and care strategies, particularly as age, *APOE* ε4 status, and cardiovascular risk appear to affect females differently.^44^ Biological factors, including gene expression and biomarker profiles, and sociocultural influences like healthcare access and caregiving roles, may also play a role.^44^ Recent evidence of more aggressive tau pathology in females^45^ further supports the need for sex-stratified approaches in aging and dementia research.

### Racial Differences in Lifetime Risk

We also observed racial disparities in dementia and MCI risk. Black adults had a higher lifetime risk of dementia compared to White adults and a higher risk of MCI before age 85, but a lower MCI risk after age 95, potentially due to factors such as younger baseline ages and higher competing mortality. Black individuals exhibited younger onset ages and higher mortality without dementia before age 85, suggesting that those who survive into their 90s may represent a healthier subgroup. This is supported by the observation that mortality without dementia after age 95 is higher among White individuals. Selective survival may therefore partially explain the reduced magnitude of racial disparities in dementia risk among the oldest-old. While our findings are consistent with the ARIC study,^12^ the magnitude of racial differences was smaller after adjusting for left truncation. This may be attributed to our cohort’s younger Black population. The racial differences in cumulative incidence of dementia was higher (44.6% in Black vs. 34.9% in White) in a Multiracial Cohort; however, these estimates were based on individuals who survived to age 90 without dementia, potentially introducing survival bias.^40^ Our results extend earlier studies ^1,13,18,38^ that focused primarily on prevalence or relative risk and did not account for competing risks. Barriers to care and potential biomarker differences may contribute to these disparities.^47–49^ Future studies should include larger and more diverse samples with extended follow-up to better characterize racial disparities in lifetime dementia and MCI risk, particularly in the oldest-old.

### Strengths and Limitations

This study has several strengths. It leveraged data from multiple large, well-characterized cohorts of community-dwelling participants without known dementia at baseline, who underwent detailed annual cognitive evaluations for up to 29 years. The annual evaluations capture rapidly progressive dementia that may exit the population in studies with intervals of 4-6 years as death from incident Alzheimer’s dementia is less than 3 years among those age 85+.^9^ The overall follow-up rate of survivors exceeds 90% reducing bias from loss to follow-up. Unlike many studies, our cohorts include a substantial number of participants over 95, allowing for more reliable lifetime risk estimates in the oldest age groups. The uniform and systematic clinical assessments across five cohorts ensure high diagnostic accuracy and consistency. We used a conservative MCI definition by requiring two consecutive assessments, which reduces misclassification and enhances diagnostic stability but may underestimate lifetime MCI risk. Another strength is the adjustment for left truncation, which minimizes bias related to staggered ages at study entry, improving accuracy compared to studies that do not account for this factor. Despite these strengths, limitations exist. The study focused primarily on Black and White adults, though Latino participants are actively being enrolled to increase diversity. We did not examine the difference in lifetime risk by genetic risk factors such as *APOE* status.

Future research should explore lifetime risk across genetic and other subgroups. Additionally, the plateauing of dementia, MCI, and death risk after age 95 is based on estimates from the competing risk models. Future studies could assess the statistical significance of this pattern using spline models or other flexible methods. Lastly, because participants join the studies voluntarily, and are older and relatively well-educated, the findings may not fully generalize to the U.S. population.

## Data Availability

Data used for this study can be requested for research purposes via the Rush Alzheimer's Disease Center Research Resource Sharing Hub.

https://www.radc.rush.edu/

## ACKNOWLEDGMENTS

The authors are grateful to the study participants and their families. They also thank the investigators and staff at the Rush Alzheimer’s Disease Center.

## STUDY FUNDING

This study is funded by National Institute on Aging Grants P30AG10161, P30AG72975, R01AG15819, R01AG34374, and R01AG022018.

## AUTHOR CONTRIBUTIONS

DAB, LY and LD contributed to the conception and design of the study; LD contributed to the acquisition and analysis of data; LD, DAB and LY contributed to the drafting of the text and preparation of figures and tables; all authors contributed to critical revision of the manuscript.

## DISCLOSURE

The authors report no relevant disclosures. Go to Neurology.org/N for full disclosures.

## Notes

### Competing Interest Statement

The authors have declared no competing interest.

### Author Declarations

The Institutional Review Board of Rush University Medical Center gave ethical approval for this work. All participants provided written informed consent.

### Summary of Updates

Replaced text to match the original June 5 submission per journal policy. No reviewer-driven changes.

