## Supplementary Files for "Lifetime risk of incident dementia and incident mild cognitive impairment in older adults"

**Supplementary Material**

Supplementary Table 1 Characteristics of Study Participants by Sex and Race in Estimating Lifetime Risk of Dementia.

| **Characteristic** | **Sex** | | **Race** | |
| --- | --- | --- | --- | --- |
|  | **Female** | **Male** | **White** | **Black** |
| Age at baseline, years | 76.53 (7.77) | 76.76 (7.58) | 78.34 (7.60) | 73.00 (6.45)* |
| Age at last visit, years | 85.50 (8.00) | 84.96 (7.48)* | 87.09 (7.45) | 82.22 (7.32)* |
| Follow up years | 8.68 (5.71) | 7.96 (5.86)* | 8.62 (5.99) | 8.61 (5.38) |
| Female, n (%) | -- | -- | 2278 (72.3) | 1007 (79.5)* |
| White, n (%) | 2278 (66.4) | 873 ( 74.0)* | -- | -- |
| Education, years | 15.51 (3.80) | 16.50 (4.29)* | 16.36 (3.82) | 15.01 (3.37)* |
| *APOE* ε4 carriers ^a^, n (%) | 754 (27.3) | 236 ( 24.9) | 595 (23.1) | 378 ( 36.5)* |
| Study, n (%) |  | * |  | * |
| ROS | 941 (27.4) | 382 ( 32.4) | 1220 (38.7) | 89 ( 7.0) |
| MAP | 1448 (42.2) | 514 ( 43.6) | 1841 (58.4) | 99 ( 7.8) |
| MARS | 575 (16.8) | 173 ( 14.7) | 0 ( 0.0) | 747 ( 59.0) |
| AA | 282 ( 8.2) | 59 ( 5.0) | 2 ( 0.1) | 330 ( 26.1) |
| LATC | 185 ( 5.4) | 52 ( 4.4) | 88 ( 2.8) | 1 ( 0.1) |
| Alive free of dementia, n (%) | 1489 (43.4) | 383 ( 32.5)* | 1022 (32.4) | 698 ( 55.1)* |
| Incident cases of dementia, n (%) | 952 (27.7) | 326 ( 27.6) | 987 (31.3) | 268 ( 21.2)* |
| Deaths without dementia, n (%) | 990 (28.9) | 471 ( 39.9)* | 1142 (36.2) | 300 ( 23.7)* |
| Age at death | 87.25 (7.50) | 85.92 (6.93)* | 88.17 (6.66) | 82.03 (7.55)* |
| Interval between age at last and at death  (median [IQR]) | 0.71 [0.41, 1.04] | 0.66 [0.35, 0.97] | 0.68 [0.37, 0.97] | 0.75 [0.42, 1.17]* |

Values are mean (SD) and for the entire analytic sample unless otherwise stated.

Abbreviations: ROS, Religious Orders Study; MAP, Memory and Aging Project; MARS, Minority Aging Research Study; AACORE, Clinical Core; LATC, Latino Core; *APOE*, Apolipoprotein E; IQR: Interquartile range

^a^ Data missing for 901 participants.

**P*<0.05.

Supplementary Table 2 Characteristics of Study Participants by Sex and Race in Estimating Lifetime Risk of MCI.

| **Characteristic** | **Sex** | | **Race** | |
| --- | --- | --- | --- | --- |
|  | **Female** | **Male** | **White** | **Black** |
| Age at baseline, years | 75.97 (7.59) | 75.57 (7.34) | 77.55 (7.50) | 72.48 (6.13)* |
| Age at last visit, years | 85.27 (8.00) | 84.52 (7.65)* | 86.85 (7.56) | 81.94 (7.19)* |
| Follow up years | 8.99 (5.75) | 8.67 (6.05) | 9.16 (6.10) | 8.82 (5.31) |
| Female, n (%) | -- | -- | 1963 (73.8) | 873 (80.8)* |
| White, n (%) | 1963 (66.2) | 698 ( 73.6)* | -- | -- |
| Education, years | 15.59 (3.81) | 16.57 (4.39)* | 16.45 (3.85) | 15.09 (3.32)* |
| *APOE* ε4 carriers ^a^, n (%) | 624 (26.3) | 171 ( 22.6) | 465 (21.5) | 315 ( 36.0)* |
| Study, n (%) |  | * |  | * |
| ROS | 821 (27.7) | 323 ( 34.1) | 1071 (40.2) | 60 ( 5.6) |
| MAP | 1214 (40.9) | 386 ( 40.7) | 1510 (56.7) | 72 ( 6.7) |
| MARS | 508 (17.1) | 141 ( 14.9) | 0 ( 0.0) | 648 ( 60.0) |
| AA | 259 ( 8.7) | 50 ( 5.3) | 2 ( 0.1) | 299 ( 27.7) |
| LATC | 165 ( 5.6) | 48 ( 5.1) | 78 ( 2.9) | 1 ( 0.1) |
| Alive free of MCI, n (%) | 1264 (42.6) | 311 ( 32.8)* | 874 (32.8) | 574 ( 53.1)* |
| Incident cases of MCI, n (%) | 1132 (38.2) | 388 ( 40.9) | 1157 (43.5) | 326 ( 30.2)* |
| Deaths without MCI, n (%) | 571 (19.2) | 249 ( 26.3)* | 630 (23.7) | 180 ( 16.7)* |
| Age at death | 86.22 (7.73) | 84.73 (7.20)* | 87.28 (6.86) | 80.88 (7.57)* |
| Interval between age at last and  at death  (median [IQR]) | 0.69 [0.41, 1.04] | 0.69 [0.38, 1.03] | 0.66 [0.37, 0.98] | 0.78 [0.48, 1.21]* |

Values are mean (SD) and for the entire analytic sample unless otherwise stated.

Abbreviations: MCI, Mild Cognitive Impairment; ROS, Religious Orders Study; MAP, Memory and Aging Project; MARS, Minority Aging Research Study; AACORE, Clinical Core; LATC, Latino Core; *APOE*, Apolipoprotein E; IQR: Interquartile range

^a^ Data missing for 790 participants.

**P*<0.05.

Supplementary Table 3 The cumulative incidence of dementia or MCI at each year of age, overall, by sex and race

| **Cumulative incidence** | **Overall** | **Sex** | | **Race** | |
| --- | --- | --- | --- | --- | --- |
|  |  | **Female** | **Male** | **White** | **Black** |
| Cumulative incidence of Dementia |  |  |  |  |  |
| Age 75 years | 6 (4,8) | 6 (4,9) | 5 (2,7) | 6 (2,9) | 6 (3,8) |
| Age 85 years | 20 (17,22) | 19 (16,22) | 22 (18,25) | 20 (16,24) | 20 (17,23) |
| Age 95 years | 39 (34,42) | 39 (35,44) | 36 (32,40) | 39 (33,45) | 41 (36,46) |
| Age 105 years | 43 (38,47) | 45 (40,50) | 39 (35,43) | 44 (37,50) | 45 (40,50) |
| Cumulative incidence of MCI |  |  |  |  |  |
| Age 75 years | 13 (11,15) | 13 (10,16) | 13 (9,17) | 10 (7,12) | 16 (12,20) |
| Age 85 years | 36 (33,39) | 35 (31,39) | 40 (35,44) | 36 (33,38) | 37 (33,42) |
| Age 95 years | 58 (54,65) | 59 (54,64) | 57 (52,62) | 60 (58,63) | 58 (53,63) |
| Age 105 years | 62 (57,67) | 63 (58,69) | 60 (55,64) | 65 (62,67) | 60 (54,65) |

Estimates are reported as percentages and indicate the cumulative incidence at the age of last observation (up to age 105 years) after accounting for the competing risk of death. The 95% CIs are reported in parentheses.

Supplementary Table 4 The cumulative incidence of death without dementia or MCI at each year of age, overall, by sex and race

| **Cumulative incidence** | **Overall** | **Sex** | | **Race** | |
| --- | --- | --- | --- | --- | --- |
|  |  | **Female** | **Male** | **White** | **Black** |
| Cumulative incidence of Death without Dementia |  |  |  |  |  |
| Age 75 years | 16 (8,24) | 15 (7,24) | 15 (11,20) | 13 (2,24) | 16 (10,21) |
| Age 85 years | 32 (26,39) | 30 (22,37) | 36 (31,41) | 29 (20,39) | 33 (28,38) |
| Age 95 years | 51 (47,57) | 49 (43,55) | 57 (53,61) | 50 (43,57) | 50 (45,55) |
| Age 105 years | 57 (53,62) | 55 (50,60) | 61 (57,65) | 56 (50,63) | 55 (50,60) |
| Cumulative incidence of death without of MCI |  |  |  |  |  |
| Age 75 years | 12 (6,18) | 11 (4,18) | 12 (8,17) | 6 (4,9) | 13 (7,18) |
| Age 85 years | 23 (17,28) | 21 (15,27) | 26 (21,30) | 18 (15,21) | 24 (19,29) |
| Age 95 years | 35 (30,40) | 33 (27,38) | 38 (34,43) | 32 (29,63) | 36 (30,41) |
| Age 105 years | 38 (33,43) | 37 (31,42) | 40 (36,45) | 35 (33,38) | 39 (33,44) |

Estimates are reported as percentages and indicate the cumulative incidence at the age of last observation (up to age 105 years). The 95% CIs are reported in parentheses.

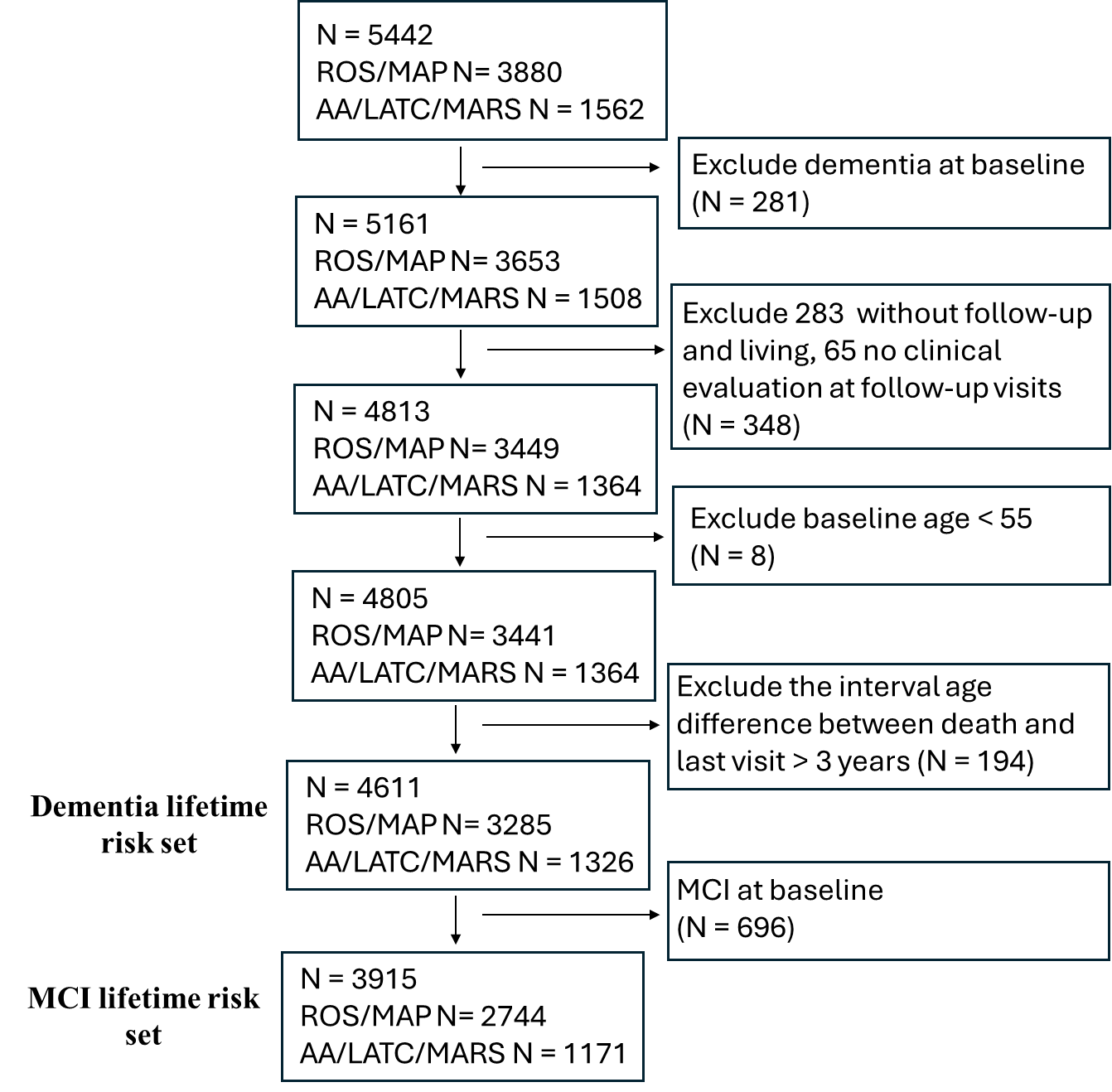

**Supplementary Figure 1. STROBE Flow Diagram.**

Abbreviations: ROS, Religious Orders Study; MAP, Memory and Aging Project; MARS, Minority Aging Research Study; AACORE, Clinical Core; LATC, Latino Core; MCI, Mild Cognitive Impairment.

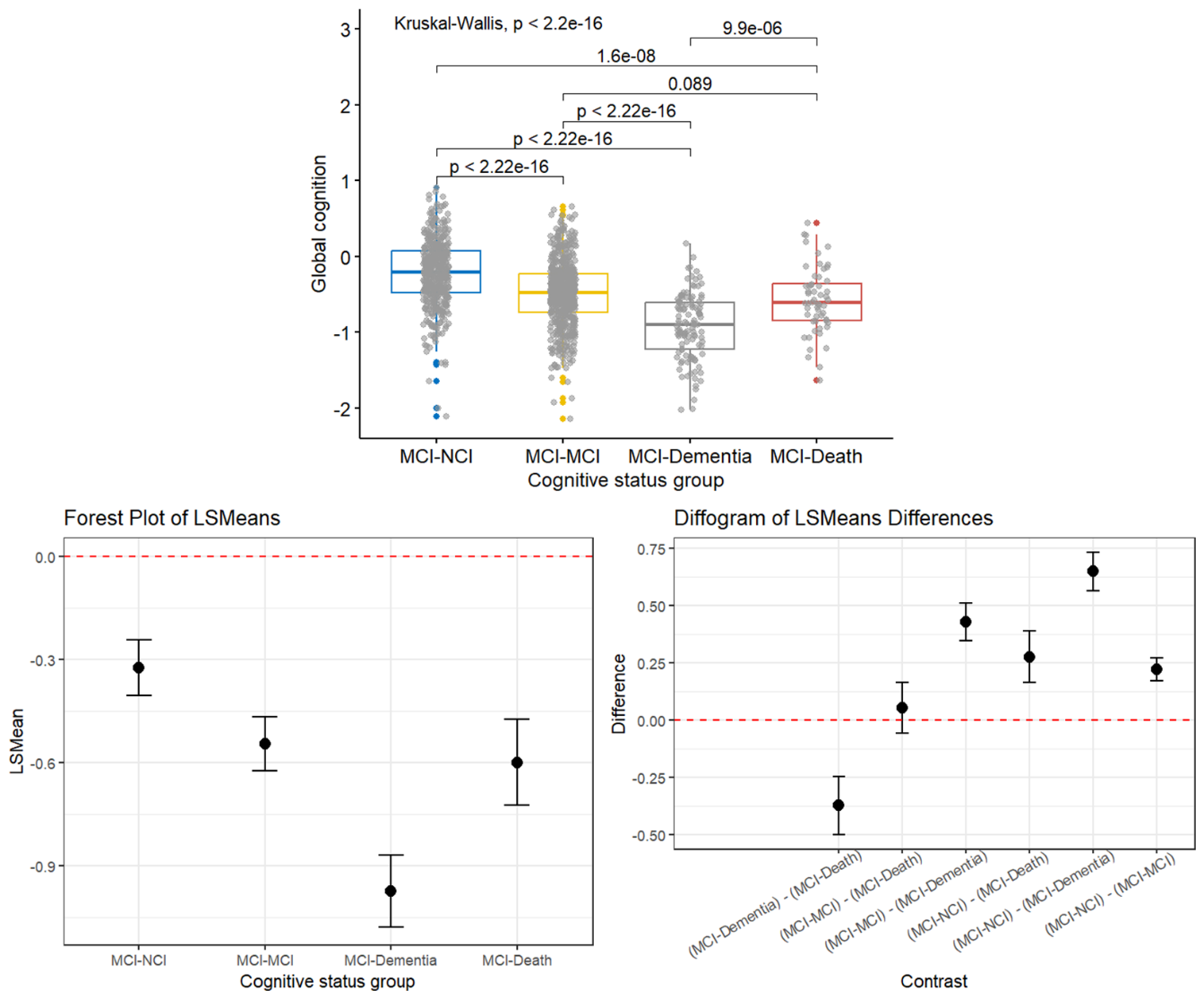

**Supplementary Figure 2. Global Cognitive Performance Across Participant Subgroups Diagnosed with MCI at Baseline**

Top panel: Boxplot showing the distribution of baseline global cognitive scores across four mild cognitive impairment (MCI) subgroups based on clinical outcomes: MCI-NCI (n = 441; reverted to no cognitive impairment at the second visit), MCI-MCI (n = 559; consistent MCI), MCI-Dementia (n = 113; progressed to dementia at the second visit), and MCI-Death (n = 56; died after the first MCI diagnosis). The Kruskal-Wallis test revealed a significant overall group difference (p < 2.2 × 10⁻¹⁶). Pairwise post-hoc comparisons showed significant differences across all groups, except between MCI-MCI and MCI-Death (annotated p-values). Bottom left: Forest plot of least square means (LSMeans) of global cognitive scores for each cognitive status group, displaying estimated means and 95% confidence intervals from a linear model adjusted for age, sex, race & ethnicity and education. Bottom right: Diffogram illustrating pairwise LSMeans differences between groups, with 95% confidence intervals. Statistically significant differences were observed among all group comparisons except between MCI-MCI and MCI-Death.

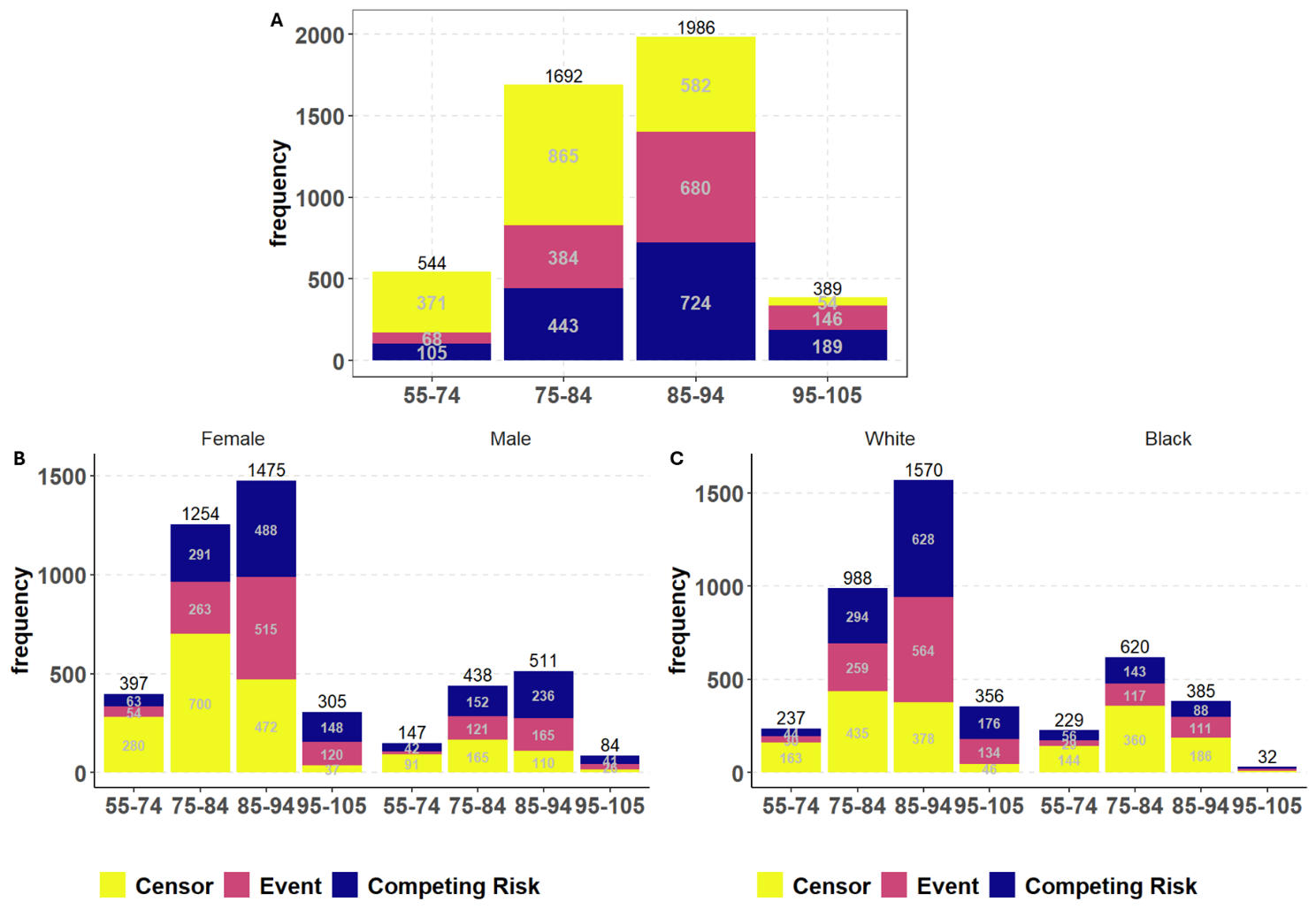

**Supplementary Figure 3. Total number of participants in the dementia lifetime risk set per decade overall, by sex, and by race (n = 4611).** Panel A shows the overall distribution, Panel B shows the distribution by sex, and Panel C shows the distribution by race. The total number of participants (black text) at each age group is displayed on top of each bar. The numbers for each outcome (censor, event, and competing risk) (gray text) are displayed within each bar. *Censor* indicates participants who were alive without dementia at the end of follow-up. *Event* indicates participants who developed incident dementia. *Competing risk* indicates participants who died without developing dementia.

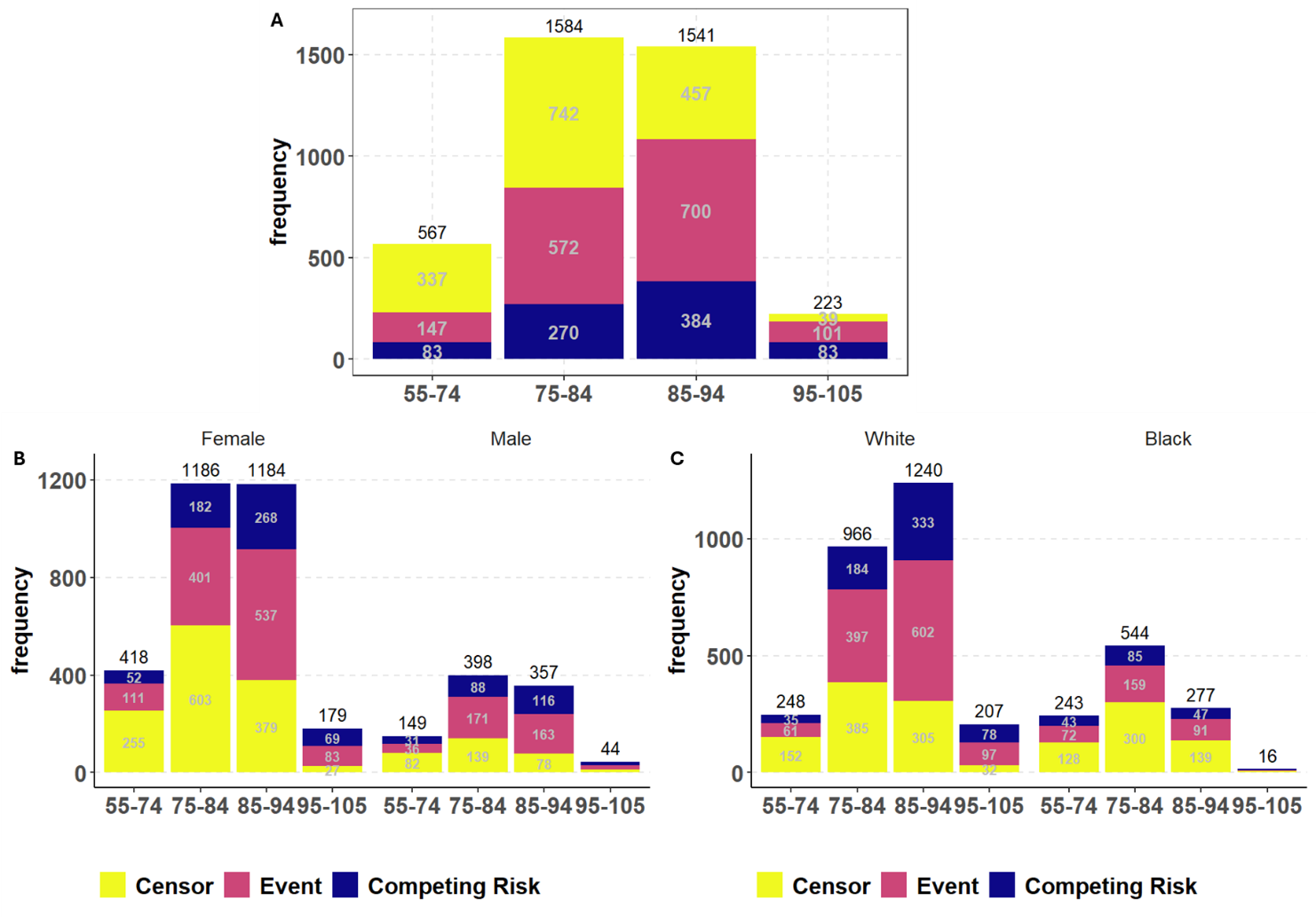

**Supplementary Figure 4. Total number of participants in the MCI lifetime risk set per decade overall, by sex, and by race (n = 3915).** Panel A shows the overall distribution, Panel B shows the distribution by sex, and Panel C shows the distribution by race. The total number of participants (black text) at each age group is displayed on top of each bar. The numbers for each outcome (censor, event, and competing risk) (gray text) are displayed within each bar. *Censor* indicates participants who were alive without MCI at the end of follow-up. *Event* indicates participants who developed incident MCI. *Competing risk* indicates participants who died without developing MCI.
